# The Potential Role of Extracellular Vesicles in COVID-19 Associated Endothelial injury and Pro-inflammation

**DOI:** 10.1101/2020.08.27.20182808

**Authors:** Balaji Krishnamachary, Christine Cook, Leslie Spikes, Prabhakar Chalise, Navneet K. Dhillon

**Author notes:** Authors contributed equally (Balaji Krishnamachary, Christine Cook). Correspondence and requests for reprints should be addressed to Navneet K. Dhillon, Division of Pulmonary and Critical Care Medicine, Department of Medicine, Mail Stop 3007, University of Kansas Medical Center, 3901 Rainbow Blvd, Kansas City, KS 66160.

## Abstract

COVID-19 infection caused by the novel severe acute respiratory syndrome coronavirus-2 (SARS-CoV-2) has resulted in a global pandemic with the number of deaths growing exponentially. Early evidence points to significant endothelial dysfunction, micro-thromboses, pro-inflammation as well as a dysregulated immune response in the pathogenesis of this disease. In this study, we analyzed the cargo of EVs isolated from the plasma of patients with COVID-19 for the identifiction of potential biomarkers of disease severity and to explore their role in disease pathogenesis. Plasma-derived EVs were isolated from 53 hospitalized patients with COVID infection and compared according to the severity of the disease. Analysis of inflammatory and cardiovascular protein cargo of large EVs revealed significantly differentially expressed proteins for each disease sub-group. Notably, members of the TNF superfamily and IL-6 family were up-regulated in patients on oxygen support with severe and moderate disease. EVs from the severe group were also enhanced with pro-thrombotic/endothelial injury factors (TF, t-PA, vWF) and proteins associated with cardiovascular pathology (MB, PRSS8, REN, HGF). Significantly higher levels of TF, CD163, and EN-RAGE were observed in EVs from severe patients when compared to patients with a moderate disease requiring supplemental O_2_. Importantly, we also observed increased caspase 3/7 activity and decreased cell survival in human pulmonary microvascular endothelial cells exposed to EVs from the plasma of patients with severe disease compared to asymptomatic group. In conclusion, our findings indicate alterations in pro-inflammatory, coagulopathy, and endothelial injury protein cargo in large EVs in response to SARS-CoV-2 infection that may be a causative agent in severe illness.

## INTRODUCTION

In December 2019, multiple cases of a pneumonia-like illness of unknown etiology were reported in Hubei Province, China. These cases were later attributed to a novel coronavirus and linked epidemiologically to a seafood market in the city of Wuhan (1). This novel coronavirus, now identified pathogenically as severe acute respiratory syndrome coronavirus-2 (SARS-CoV-2), has lead to an ongoing global pandemic impacting six continents and has been linked to more than 819,609 deaths as of June 26, 2020 (2).

There is robust early evidence that infection with SARS-CoV-2 (COVID-19) is associated with significant endothelial dysfunction and a dysregulated immune response. Like the SARS-CoV pathogen that led to epidemics in 2002 and 2003, SARS-CoV-2 enters cells via the binding of its spike protein to angiotensin converting enzyme 2 (ACE2) receptors. ACE2 receptors are abundantly present in pulmonary alveolar type II and endothelial cells, thereby making the lungs and pulmonary vasculature susceptible to SARS-CoV-2-induced inflammation and injury (3).Further, alveolar capillary micro-thrombi and endothelial damage with evidence of intracellular virus have been noted on post-mortem analysis of infected lungs (4).

While there is growing evidence that endothelial injury, vascular remodeling, and coagulopathy are key consequences of COVID-19 infection, it remains unclear how the virus induces these changes. Extracellular vesicles (EVs) carry coding and non-coding RNA, proteins, DNA fragments, and lipids, which facilitate crosstalk between cells. This transfer of EV cargo plays a significant role in a number of diseases processes, including cardiovascular disease, pulmonary hypertension, and various malignancies (5). EVs can be released in response to thrombin, shear stress, complement activation, and inflammation, among other pathophysiologic triggers. In this study, we analyzed the alterations in the plasma-derived EVs from patients infected with SARS-CoV-2 to further investigate the role(s) EVs play in COVID-19 pathophysiology.

## MATERIALS AND METHODS

### Human samples and data collection

Blood samples (EDTA plasma, whole blood, and serum) were collected from 53 hospitalized subjects 18 years of age or older with a confirmed diagnosis of COVID-19. All subjects were enrolled in the University of Kansas Health System’s (TUKHS) COVID-19 Biorepository. As participants in the biorepository, each subject had blood samples collected between 3 and 7 a.m. every three days from time of enrollment until time of discharge or until determined to be COVID-recovered according to institutional Infection Prevention and Control guidelines. Only baseline EDTA plasma samples were used in this analysis. Demographic information and clinical data were collected and stored in a secure database. Vital signs, oxygen and life support needs, and laboratory data were used to calculate acute physiology and early warning scores for patients on lab draw days. Written informed consent for participation in the Biorepository was obtained from all patients or their surrogates. Archived EDTA plasma samples from five healthy controls were included for comparison. This study and the COVID-19 Biorepository were approved by the University of Kansas Medical Center Institutional Review Board.

A confirmed case of COVID-19 infection was defined as a patient with a positive SARS-CoV-2 nucleic acid by RT-PCR. All but one patient tested positive on a nasopharyngeal swab. One patient was presumed positive based on symptoms and reactive serology. Patients were classified by World Health Organization Clinical Progression Scale score (7). Specifically, they were grouped by peak score during their hospitalization. Patients were considered asymptomatic if they had a positive SARS-CoV-2 RT-PCR but no COVID-related respiratory symptoms and were hospitalized for another medical reason (Asymptomatic group). These patients were tested for COVID-19 as part of a hospital-wide admission or pre-operative mandate. The remaining patients were categorized as having moderate disease with no oxygen requirement (Moderate-No O_2_), moderate disease requiring supplemental oxygen by simple face mask or nasal prongs (Moderate-On O_2_), or severe disease requiring oxygen delivery by non-rebreather mask, non-invasive ventilation, or heated high flow nasal cannula at a minimum (Severe group). Uninfected healthy EDTA plasma collected before COVID-19 pandemic was also used for comparison (Healthy group, N=6). Several clinical laboratory parameters were checked, every 1-3 days, as part of center-specific standard care for hospitalized COVID-19-infected patients. These included white blood cell (WBC) count, absolute lymphocyte count, creatinine, lactate dehydrogenase (LDH), ferritin, D-dimer, and C-reactive protein (CRP).

### Isolation of 20K and 100k pellet particles from plasma samples

About 1ml of EDTA plasma samples from different groups were centrifuged at 2500g for 15 minutes at room temperature to obtain the platelet free plasma (PFP). The PFP was further centrifuged at 20,000g for 15 minutes at 4°C to isolate the 20K pellet large vesicles. The 20K pellet was further washed with PBS and centrifuged again at 20,000g for 15 minutes at 4°C followed by re-suspension in 200µl of PBS. The plasma supernatant after the first 20,000g spin was filtered through 0.22µm filter and subjected to ultracentrifugation at 100,000g for 70 minutes at 4°C. The pellet was washed with PBS and centrifuged again at 100,000g for 70 minutes at 4°C and finally re-suspended in approximately 500µl of PBS. To determine the total number of particles and mean particle size, Nanosight analysis was performed on both 20K and 100K pellets (Malvern Panalytical, UK).

### Transmission electron microscopy

A few microliters of 20K and 100K pellet suspended in PBS was placed on the carbon coated copper grid, followed by 1% uranyl acetate staining to enhance the contrast. The images were acquired using the Transmission electron microscopy at 100KV emission (Jeol, USA).

### Caspase 3/7 Assay

Primary human pulmonary microvascular endothelial cells (HPMECs) were plated on a 96-well plate (Corning USA) in complete endothelial cell growth medium (ScienCell, USA). After 24h, the media was removed, and the cells were washed followed by treatment with 3µg of 20K pellet EVs in serum free endothelial cell media. After 24h, caspase 3/7 assay was performed using Cell Meter™ Caspase 3/7 Activity Apoptosis Assay Kit, according to the manufacturer’s instructions (AAT Bioquest, USA).

### MTS Assay

For survival assays, HPMECs were plated on 96-well plates in complete endothelial cell growth medium and after 24h, the cells were treated with 3ug of EVs in 0.5% FBS containing media. After 48h, MTS assay was performed using CellTiter 96® Aqueous One Solution Cell Proliferation Assay kit (Promega, Madison, WI).

### Proximity Extension analysis (PEA)

The 20K pellet cargo was analyzed for inflammatory and cardiovascular markers using Olink inflammatory and Cardiovascular II and III panels (Olink, Boston, USA). The results were expressed as normalized protein expression (NPX) expressed on a log2 scale, and a high NPX value indicates higher protein expression.

### Statistical Analyses

The demographic and clinical characteristics for the four disease status groups were summarized with mean and standard deviation for the continuous variables and with frequencies and percentages for categorical variables, **Table 1**. The differences in the continuous variables across the groups were assessed using analysis of variance (ANOVA) followed by post hoc comparisons using Tukey’s test. Normality assumption of the data were examined by histogram and boxplots prior to performing the ANOVA analyses. Log transformation was used for the outcomes not satisfying the normality assumption. The categorical outcomes across the groups were assessed using Fisher exact test. The differences in the protein expressions among the five groups (Healthy, Asymptomatic, Moderate-No O_2_, Moderate-On O_2_, and Severe) were assessed using ANOVA analyses followed by pairwise post hoc comparison using Tukey’s test. Spearman’s correlation analyses were carried out to assess the relationship between each protein with age, BMI, WBC count, LDH, ferritin, CRP, creatinine, D-dimer, and absolute lymphocyte count, for all subjects or symptomatic patients and then by separate groups. All the tests were considered statistically significant if p-values were less than 0.05. The statistical analyses were carried out using the statistical software R version 4.0.0(R Code Team, 2020). For the Nanosight and cell-culture experiments, statistical analyses were carried out using Graph Pad Prism 8. One-way ANOVA was performed among the groups followed by Bonferroni’s multiple comparison test.

**Table 1:**
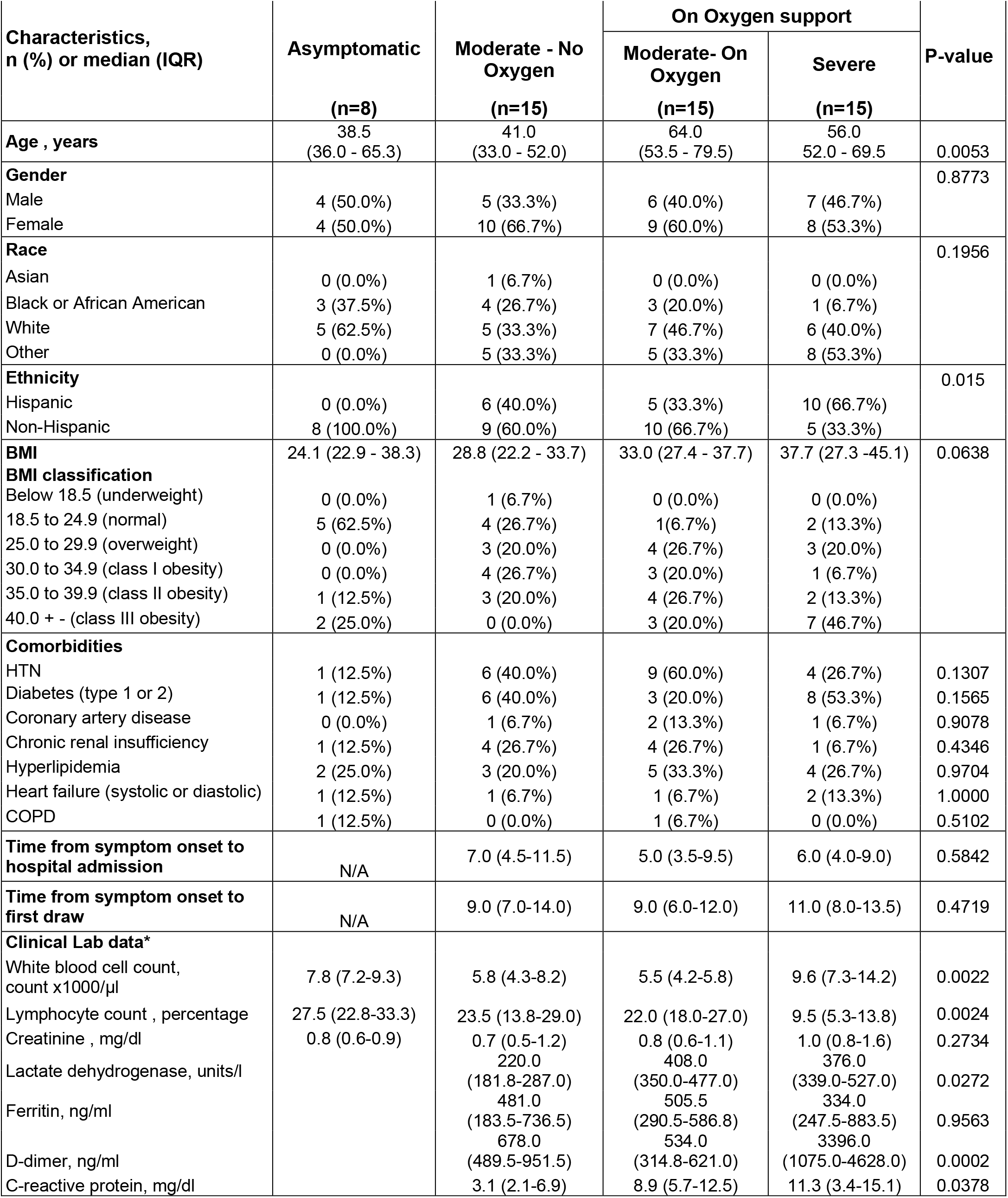
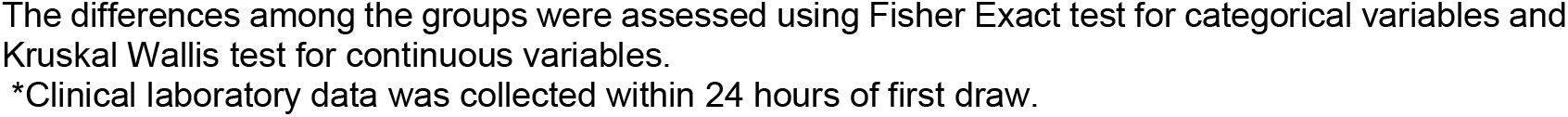
Demographics and characteristics of patients included in analysis.

| Characteristics,<br>n (%) or median (IQR) | Asymptomatic<br>(n=8) | Moderate - No<br>Oxygen<br>(n=15) | On Oxygen support |  | P-value |
| --- | --- | --- | --- | --- | --- |
|  |  |  | Moderate- On<br>Oxygen<br>(n=15) | Severe<br>(n=15) |  |
| **Age , years** | 38.5<br>(36.0 - 65.3) | 41.0<br>(33.0 - 52.0) | 64.0<br>(53.5 - 79.5) | 56.0<br>52.0 - 69.5 | 0.0053 |
| **Gender** |  |  |  |  | 0.8773 |
| Male | 4 (50.0%) | 5 (33.3%) | 6 (40.0%) | 7 (46.7%) |  |
| Female | 4 (50.0%) | 10 (66.7%) | 9 (60.0%) | 8 (53.3%) |  |
| **Race** |  |  |  |  | 0.1956 |
| Asian | 0 (0.0%) | 1 (6.7%) | 0 (0.0%) | 0 (0.0%) |  |
| Black or African American | 3 (37.5%) | 4 (26.7%) | 3 (20.0%) | 1 (6.7%) |  |
| White | 5 (62.5%) | 5 (33.3%) | 7 (46.7%) | 6 (40.0%) |  |
| Other | 0 (0.0%) | 5 (33.3%) | 5 (33.3%) | 8 (53.3%) |  |
| **Ethnicity** |  |  |  |  | 0.015 |
| Hispanic | 0 (0.0%) | 6 (40.0%) | 5 (33.3%) | 10 (66.7%) |  |
| Non-Hispanic | 8 (100.0%) | 9 (60.0%) | 10 (66.7%) | 5 (33.3%) |  |
| **BMI** | 24.1 (22.9 - 38.3) | 28.8 (22.2 - 33.7) | 33.0 (27.4 - 37.7) | 37.7 (27.3 -45.1) | 0.0638 |
| **BMI classification** |  |  |  |  |  |
| Below 18.5 (underweight) | 0 (0.0%) | 1 (6.7%) | 0 (0.0%) | 0 (0.0%) |  |
| 18.5 to 24.9 (normal) | 5 (62.5%) | 4 (26.7%) | 1(6.7%) | 2 (13.3%) |  |
| 25.0 to 29.9 (overweight) | 0 (0.0%) | 3 (20.0%) | 4 (26.7%) | 3 (20.0%) |  |
| 30.0 to 34.9 (class I obesity) | 0 (0.0%) | 4 (26.7%) | 3 (20.0%) | 1 (6.7%) |  |
| 35.0 to 39.9 (class II obesity) | 1 (12.5%) | 3 (20.0%) | 4 (26.7%) | 2 (13.3%) |  |
| 40.0 + - (class III obesity) | 2 (25.0%) | 0 (0.0%) | 3 (20.0%) | 7 (46.7%) |  |
| **Comorbidities** |  |  |  |  |  |
| HTN | 1 (12.5%) | 6 (40.0%) | 9 (60.0%) | 4 (26.7%) | 0.1307 |
| Diabetes (type 1 or 2) | 1 (12.5%) | 6 (40.0%) | 3 (20.0%) | 8 (53.3%) | 0.1565 |
| Coronary artery disease | 0 (0.0%) | 1 (6.7%) | 2 (13.3%) | 1 (6.7%) | 0.9078 |
| Chronic renal insufficiency | 1 (12.5%) | 4 (26.7%) | 4 (26.7%) | 1 (6.7%) | 0.4346 |
| Hyperlipidemia | 2 (25.0%) | 3 (20.0%) | 5 (33.3%) | 4 (26.7%) | 0.9704 |
| Heart failure (systolic or diastolic) | 1 (12.5%) | 1 (6.7%) | 1 (6.7%) | 2 (13.3%) | 1.0000 |
| COPD | 1 (12.5%) | 0 (0.0%) | 1 (6.7%) | 0 (0.0%) | 0.5102 |
| **Time from symptom onset to**<br>**hospital admission** | N/A | 7.0 (4.5-11.5) | 5.0 (3.5-9.5) | 6.0 (4.0-9.0) | 0.5842 |
| **Time from symptom onset to**<br>**first draw** | N/A | 9.0 (7.0-14.0) | 9.0 (6.0-12.0) | 11.0 (8.0-13.5) | 0.4719 |
| **Clinical Lab data\*** |  |  |  |  |  |
| White blood cell count,<br>count x1000/μl | 7.8 (7.2-9.3) | 5.8 (4.3-8.2) | 5.5 (4.2-5.8) | 9.6 (7.3-14.2) | 0.0022 |
| Lymphocyte count , percentage | 27.5 (22.8-33.3) | 23.5 (13.8-29.0) | 22.0 (18.0-27.0) | 9.5 (5.3-13.8) | 0.0024 |
| Creatinine , mg/dl | 0.8 (0.6-0.9) | 0.7 (0.5-1.2) | 0.8 (0.6-1.1) | 1.0 (0.8-1.6) | 0.2734 |
| Lactate dehydrogenase, units/l |  | 220.0<br>(181.8-287.0) | 408.0<br>(350.0-477.0) | 376.0<br>(339.0-527.0) | 0.0272 |
| Ferritin, ng/ml |  | 481.0<br>(183.5-736.5) | 505.5<br>(290.5-586.8) | 334.0<br>(247.5-883.5) | 0.9563 |
| D-dimer, ng/ml |  | 678.0<br>(489.5-951.5) | 534.0<br>(314.8-621.0) | 3396.0<br>(1075.0-4628.0) | 0.0002 |
| C-reactive protein, mg/dl |  | 3.1 (2.1-6.9) | 8.9 (5.7-12.5) | 11.3 (3.4-15.1) | 0.0378 |

The differences among the groups were assessed using Fisher Exact test for categorical variables and Kruskal Wallis test for continuous variables.
\*Clinical laboratory data was collected within 24 hours of first draw.

## RESULTS

### Demographic and clinical characteristics

Of the 53 hospitalized patients with blood samples included in our analysis, 9 had no respiratory symptoms (Asymptomatic), 15 had moderate disease but were not hypoxic (Moderate-No O_2_), 15 had moderate disease and were hypoxic (Moderate-On O_2_), and 15 had severe COVID-19 infection (Severe) (Table 1). Although mean age of patients on O_2_ support was higher compared to other groups, no difference was observed between patients in ‘Moderate-On O_2_’ and ‘Severe’ groups. Mean time from symptom onset to hospital admission was 8 days. Mean time from symptom onset to baseline lab draw was 11 days with no statistical differences across the groups. Forty-one percent of patients were male and more than one-third of patients included in the study were identified as Hispanic. Although nearly one-half of patients met criteria for class I, II, or III obesity, the difference in the average body mass index (BMI) of patients across the groups was not significant. The most common comorbidities among these patients were hypertension (38%), diabetes mellitus (type 1 or 2) (34%), hyperlipidemia (26%), and chronic renal insufficiency (18.9%). Thirty percent of patients were current or former smokers. Sixty-six percent of patients had no history of tobacco, alcohol, or drug abuse.

Among asymptomatic patients included in our analysis, reasons for hospitalization included post-operative *Clostridium difficile* infection, hyperemesis gravidarum, sickle cell pain crisis, acute variceal hemorrhage, heart failure exacerbation with newly reduced left ventricular ejection fraction, post-operative pelvic abscess, gunshot wound, and osteomyelitis. Of the 15 patients with severe disease, six were mechanically ventilated, and five died during hospitalization.

Regarding laboratory data collected within 24 hours of first lab draw from symptomatic COVID-19 patients, there was a significant rise in median D-dimer (p=0.0002), CRP (p=0.0378), and LDH (p=0.0272) levels according to illness severity in symptomatic COVID-19 patients(Table 1). There also was a significant increase in median WBC count in patients from the severe group compared to those in the moderate groups; however, WBC count values remained within the normal range. There was a significant drop in lymphocyte count in those with severe disease (p=0.0024).

### Characterizing the 20K and 100K pellet plasma derived EVs

20K and 100K pellet EVs isolated from the plasma of all SARS-CoV-2 positive patients and healthy uninfected controls were compared for total number and size using nanosight analysis. In the 20K pellet derived large EVs (LEVs), a significant increase in the total particle number and mean particle size was observed in the Moderate-No O_2_ group when compared to the Asymptomatic and Moderate-On O_2_ groups (Fig. 1A-B). However, a significant increase in the total particle number and mean particle size of 100K pellet small EVs (SEVs) was observed in the Severe group when compared to the Asymptomatic and Moderate-On O_2_ groups. As expected, the mean particle size was higher in LEVs when compared to the SEVs. This was further confirmed by TEM analysis. As shown in the represented TEM images (Fig. 1C), LEVs showed particles of size ranging from 100-300nm, whereas 100K SEVs showed smaller particles in the range of 30-100nm.

**Figure 1:**
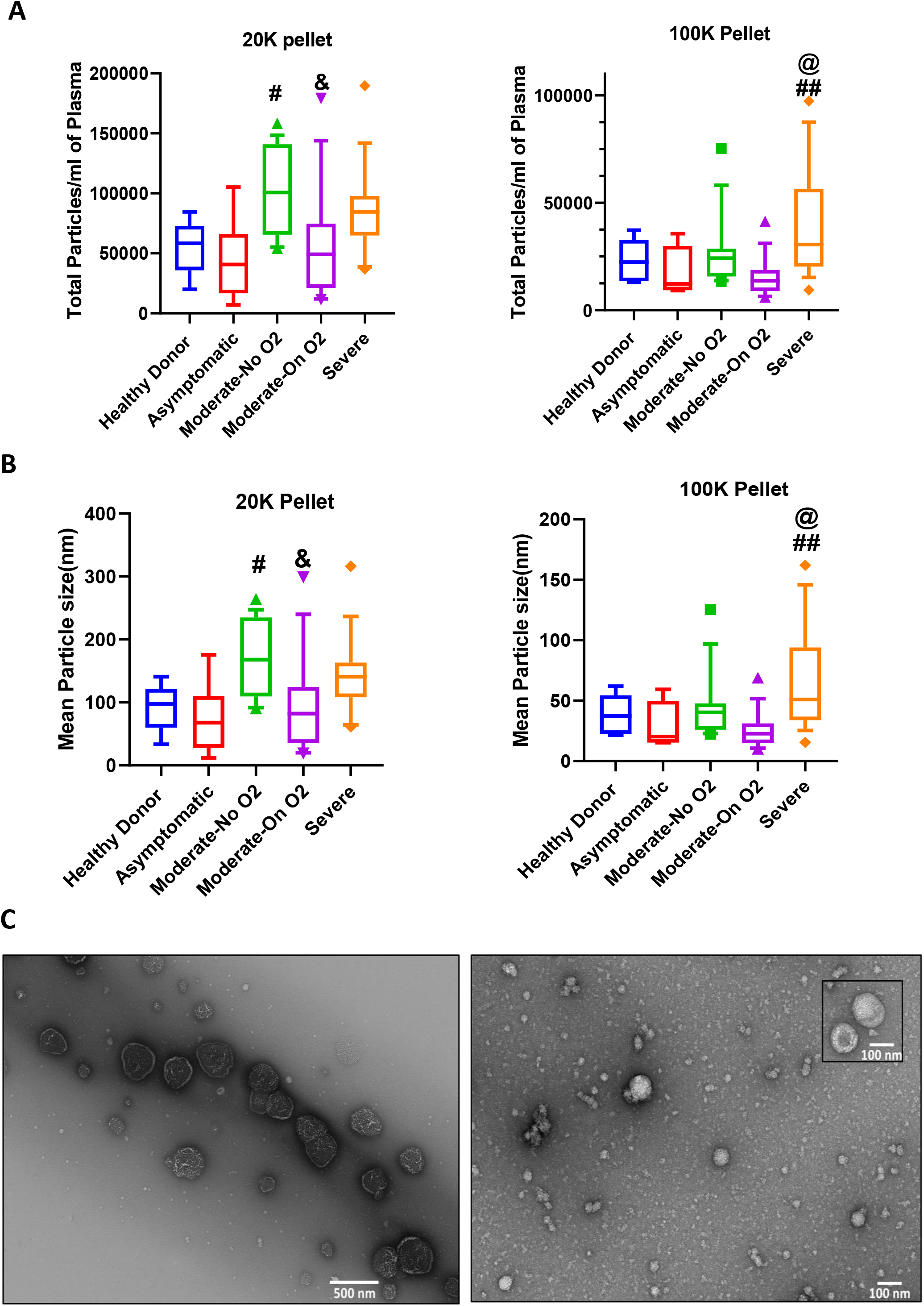
Characterization of extracellular vesicles isolated from plasma of COVID19-patients and healthy individuals. Extracellular vesicles (EVs) were isolated from 1ml of EDTA-plasma of healthy controls (n=6), asymptomatic (n=8), Moderate-No O_2_ (n=15), Moderate-On O_2_ (n=15), Severe (n=15) by centrifugation at 20,000g (20K pellet large EVs) and at 100,000g (100K pellet small EVs). **A-B)** Nano sight analysis shows total number of particles **(A)** and mean particle size **(B)**. Boxes in A-B panel depicts median and IQR and whiskers represent 10-90 percentiles. **C)** Characterization of 20K pellet and 100K pellet by TEM. Representative TEM image of 20K pellet (left panel) at 10,000X and 100K pellet (right panel) at 25,000X magnification. The insert represents a different image of the same 100K pellet sample at same magnification to show the bigger particles. # p<0.05, ##p<0.01 vs. Asymptomatic, & p<0.05 vs Moderate-No O_2_, @ p<0.05 vs. Moderate-On O_2_.

### Analysis of LEV protein cargo for inflammatory and cardiovascular biomarkers

The Olink analysis was performed on the LEVs from five different groups (Healthy, n=6, Asymptomatic n=6, Moderate-No O_2_ n=13, Moderate-On O_2_ n=14 and Severe n=9) using inflammation and cardiovascular II and III panels with each panel having the ability to detect about 92 proteins. The statistically significant inflammatory and cardiovascular proteins in LEVs from all groups are presented as a heatmap, showing the presence of pattern of protein expression according to the disease severity (Supplementary figure 1, p<0.05). Figure 2A shows the Venn diagram of the statistically significant differentially expressed (p<0.05) proteins for each of the Asymptomatic, Moderate-No O_2_, Moderate-On O_2_, and Severe subjects with respect to healthy controls. Of these, 2 were up-regulated only in ’Asymptomatic’, 4 only in ‘Moderate-No O_2_’, 10 in ‘Moderate-On O_2_’ and 14 were only in the ’Severe’ group. Some of the proteins were found to be up-regulated in multiple groups as also listed in Volcano plots (Fig. 2B). These volcano plots were created to show the differentially expressed proteins in the pairwise post hoc comparisons.

**Figure 2:**
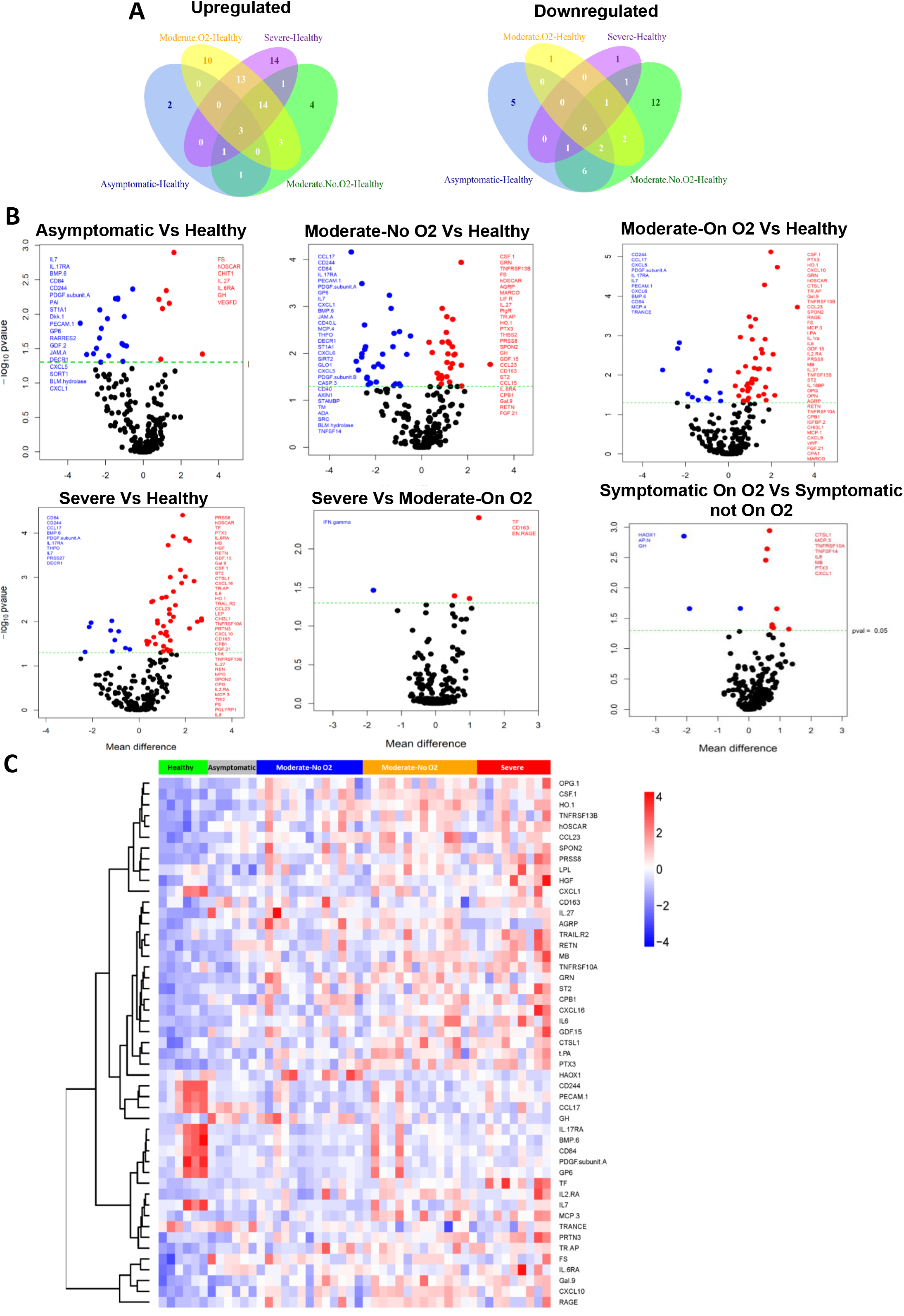
Proximity Extension Analysis (PEA) of large extracellular vesicles. 20K pellet of healthy controls (n=6), asymptomatic (n=6), Moderate-No O_2_ (n=13), Moderate-On O_2_ (n=14), Severe (n=9) were analyzed for its protein cargos by employing Olink’s multiplex PEA platform. **A)** Venn diagram represents the total number of differentially expressed up-regulated and down-regulated proteins between different groups (p<0.05). **B)** Volcano plots show pairwise post hoc comparisons of differentially expressed proteins in various groups. The plot was constructed using -log_10_ (p value) against the mean difference. The green dotted horizontal line represents p=0.05. **C)** Heat map of hierarchical clustering of proteins which were differentially expressed (p<0.01).

Interestingly, more proteins were observed to be down-regulated in patients from the Asymptomatic and Moderate-No O_2_ groups while only 1 protein each was down-regulated in the Moderate-On O_2_ and Severe groups as illustrated in Venn diagram. The Volcano plots list the down-regulated proteins in each group compared to the healthy subjects (p<0.05). Comparison of differentially expressed proteins between symptomatic patients not on O_2_ (Moderate-No O_2_) and symptomatic patients on O_2_ (Moderate-On O_2_ plus Severe groups) showed 8 up-regulated (Cathepsin L1, MCP-3, TNFRSF10A/DR4, TNFSF14, Interleukin-6, Myoglobin, Pentraxin-3 and CXCL1) and 3 down-regulated (Hydroxyacid oxidase 1, Aminopeptidase N and Growth Hormone) proteins in COVID-19 patients on external O_2_ support. Furthermore, when we compared the Severe and Moderate-On O_2_ group patients, 3 proteins, Tissue factor (TF), macrophage marker CD163, and pro-inflammatory extracellular newly identified receptor for advanced glycation end products (RAGE) binding protein (EN-RAGE) (aka S100-A12), were found to be significantly up-regulated along with down-regulation of IFN-γ (Fig. 2B).

Heatmap of differentially expressed proteins generated using p<0.01 (Fig. 2C) highlights the alterations in pro-inflammatory, thrombosis, endothelial injury, and angiogenesis-related proteins with the increase in severity of the disease. Significant changes were observed in IL-6 family cytokines in addition to IL-6, as mentioned above (Fig. 2C and Fig. 3). Increased levels of Interleukin-6 receptor subunit alpha (IL-6RA) and a IL-6 inducer, Oncostatin-M (OSM), was observed in the Severe group in comparison with the healthy subjects. IL-27 was up-regulated in all groups compared to uninfected healthy controls, while a decreased trend in leukemia inhibitory factor receptor (LIF-R) was observed in patients on oxygen support when compared to those in the Moderate-No O_2_ group (Fig. 2C). In addition, IL-8 showed maximum increase in the Severe group, while IL-18 was significantly up-regulated in both Moderate groups and the Severe group when compared to the Asymptomatic group (p<0.01). Correspondingly, IL-18R1 showed an increased trend in the Severe group when compared to all other groups with almost the same increase in IL-18 BP as other COVID-19-positive groups (Supplementary figure 1). Levels of IFN-γ were found to be higher in LEVs from the plasma of patients in the Moderate-On O_2_ group compared to Asymptomatic or symptomatic Moderate-No O_2_ groups; however, as mentioned above IFN-gamma levels were significantly lower in the Severe group (Fig. 3). IL-12B, a known inducer of IFN-γ production, was, in parallel, found to be lower in the Severe group compared to other COVID-19-positive groups.

**Figure 3:**
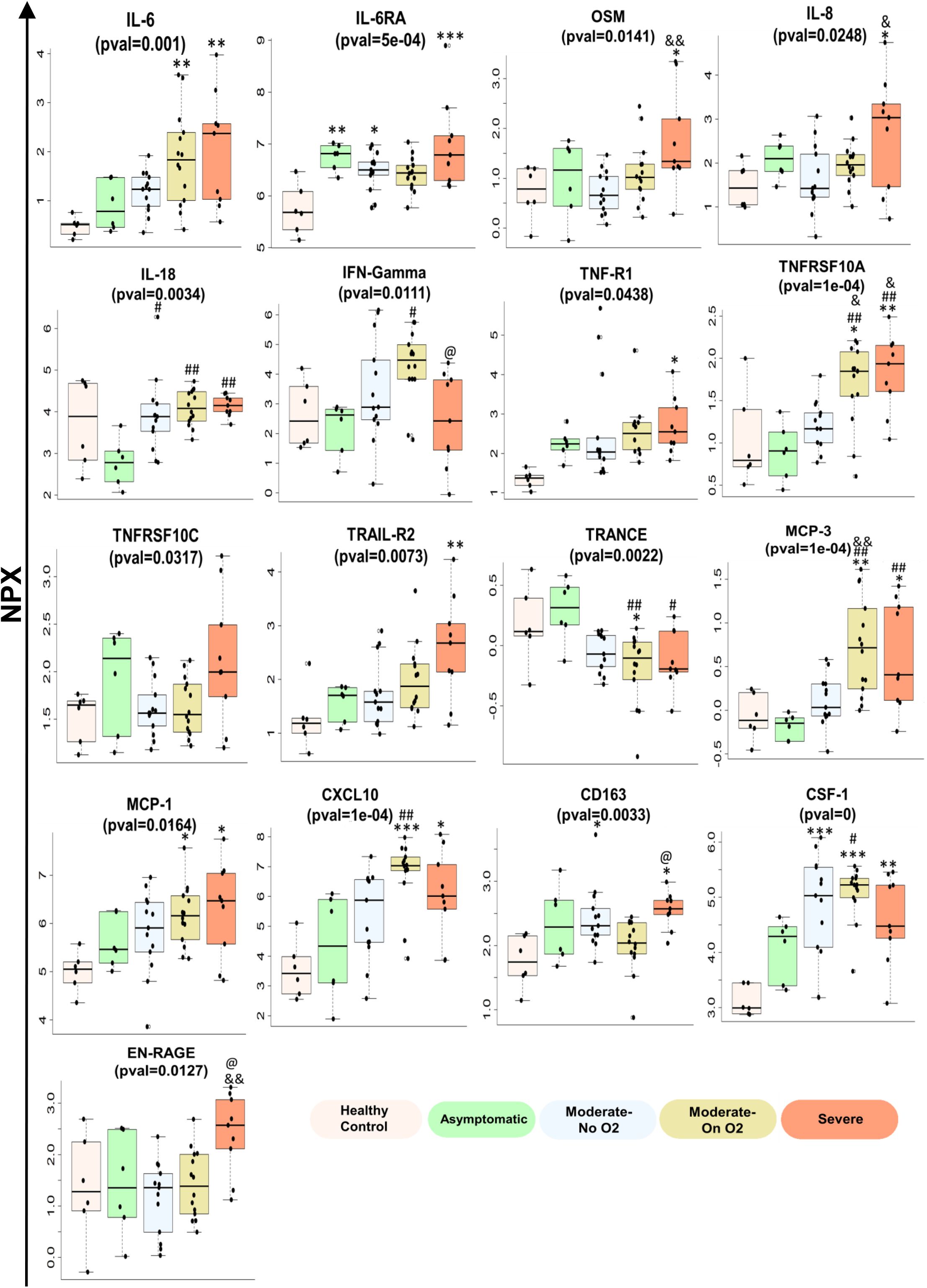
Box-whisker plots showing few of the differentially expressed inflammatory proteins in large EVs. 20K pellet EVs from different groups were analyzed for inflammatory cargo using PEA platform. Boxes span from quartile 1 and 3 with median showing in the middle and whiskers extend to 1.5 times the IQR from the box.* p<0.05, **p<0.01, ***p<0.001 vs. healthy controls, # p<0.05, ##p<0.01 vs. Asymptomatic, & p<0.05, && p<0.01 vs. Moderate-No O_2_, @ p<0.05 vs. Moderate-On O_2_.

Most importantly, LEVs from the Severe group were noted to be highly enriched in pro-inflammatory TNF-α and TNF-receptor superfamily proteins. Tumor necrosis factor receptor 1 (TNF-R1), TNF-related apoptosis-inducing ligand receptor 2 (TRAIL-R2), Tumor necrosis factor receptor superfamily member 10A (TNFRSF10A, aka TRAIL-R1 or DR4), TNFRSF10C (TRAIL-R3), Tumor necrosis factor ligand superfamily member 13B and 14 (TNFSF13B and TNFSF14), Osteoprotegerin (OPG), and Tartrate-resistant acid phosphatase type 5 (TRAP) were significantly up-regulated in patients with Moderate and/or Severe disease on O_2_ support (Figures 2C and 3). In parallel, TNF-related activation-induced cytokine (TRANCE) was significantly down-regulated in the Moderate-On O_2_ and Severe groups when compared to the Asymptomatic group (Fig. 3). Elevated levels of chemokines such as MCP-1, MCP-3, CXCL10 and CXCL16 were also found in Moderate-On O_2_ and/or Severe groups. In addition to significant high expression of CD163 in severe group EVs as mentioned above, macrophage colony stimulating factor (CSF-1) was also significantly high in both the Moderate and Severe groups when compared to the healthy group (Figures 2C and 3)

Significantly higher levels of cardiovascular disease-related proteins were also observed in 20K pellet EVs from COVID-19 patients. Thrombosis, coagulation, and endothelial injury-associated markers; tissue factor (TF), tissue plasminogen activator (t-PA), von Willebrand factor (vWF), chitinase-3-like protein 1 (CHI3L1), and renin (REN), were significantly increased in the Severe group when compared to the healthy groups, while an increased trend was observed in Chitotriosidase-1 (CHIT1) (p=0.07). Alternatively, ADAMTS13 (aka vWF-cleaving protease), an inhibitor of thrombus formation, was found to be decreased in the Severe group when compared to healthy controls (Figs. 2C and 4A).

**Figure 4:**
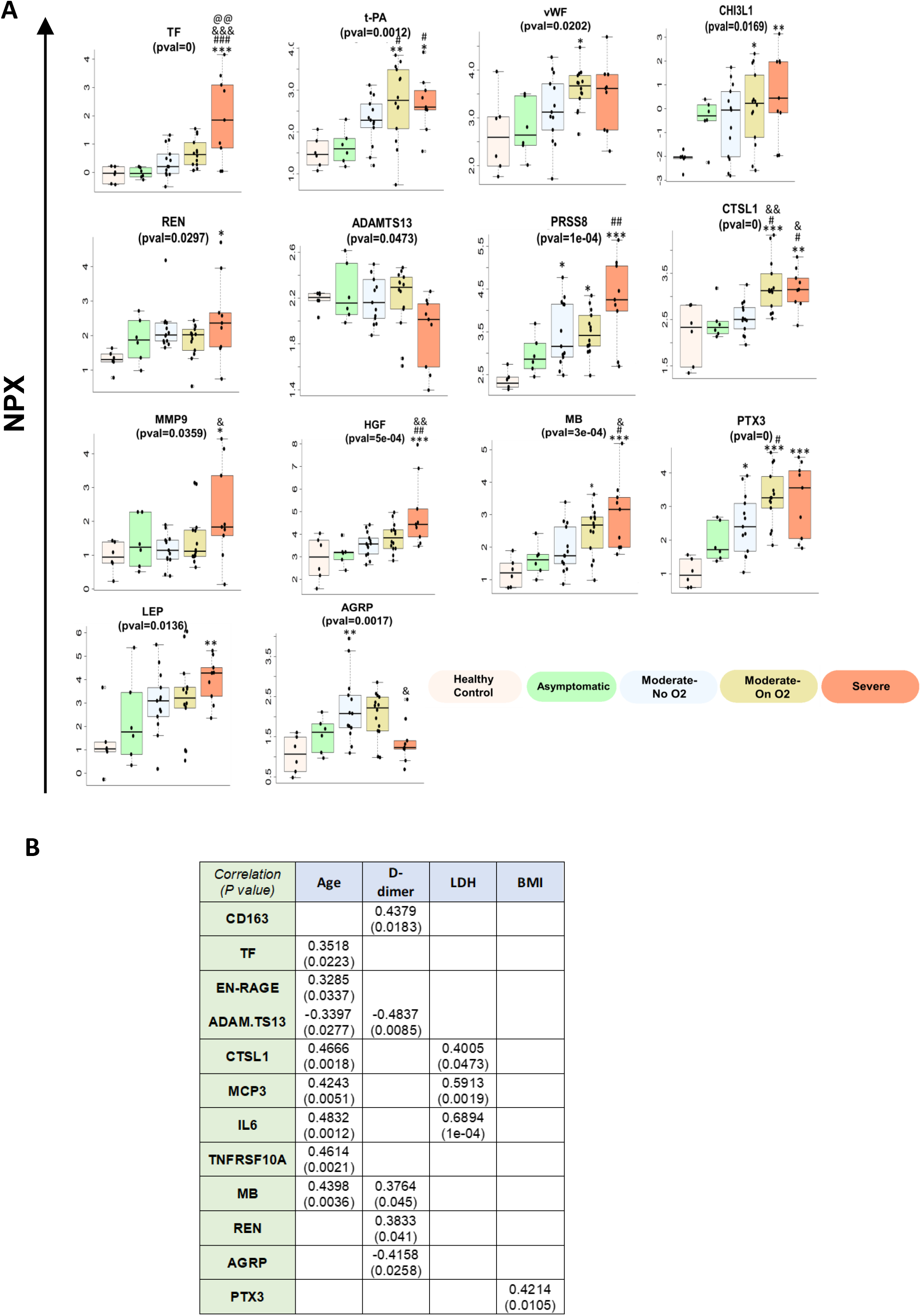
A) Differentially expressed cardio-vascular disease related proteins in large extracellular vesicles. Boxes in box plots represent median/ IQR and whiskers extend to 1.5 times the IQR from the box. * p<0.05, **p<0.01, ***p<0.001 vs. healthy controls, # p<0.05, ##p<0.01, ###p<0.001 vs. Asymptomatic, & p<0.05, && p<0.01, &&&p<0.001 vs. Moderate-No O_2_, @@ p<0.01 vs. Moderate-On O_2_. **B)** Table lists some of the significantly altered LEV proteins that significantly correlated with age and/or other clinical parameters. Spearman’s correlation analyses were carried out to assess the relationship of protein with age for all subjects and; BMI, LDH, and D-dimer for symptomatic patients.

Prostasin (PRSS8), a serine protease that regulates the epithelial sodium channel and thereby worsens airway clearance and hypertension, was identified as the most significantly up-regulated protein in the Severe group patients when compared with the healthy uninfected group (Figs. 2B and 4A). In addition, many other proteases involved in tissue remodeling, such as cathepsin L (CTSL1), carboxy peptidases (CPA1 and CPB1), and matrix metalloproteinase (MMP9) involved in tissue remodeling were at higher levels in EVs from COVID-19 patients. The potential biomarkers of cardiovascular disease, long pentraxin (PTX3) and hepatocyte growth factor (HGF) as well as myoglobin (MB), a myocyte protein and adipokine leptin (LEP) were among other important up-regulated proteins associated with 20K EVs from Severe and/or Moderate group patients (Fig. 4A).

### Correlation of significantly altered proteins in LEVs with Age, D-Dimer, LDH and BMI

For correlation regression analysis of proteins with clinical lab parameters, we included only symptomatic patients due to lack of data from most of the asymptomatic patients. From the significantly altered proteins between the Moderate-On O_2_ and Severe groups, EN-RAGE and TF showed significant correlation with age in all subjects (Fig. 4B). However, CD163 showed positive correlation with only D-Dimer in symptomatic patients. Meanwhile, ADAMTS13 (p=0.06 Severe vs. Moderate-On O_2_) was negatively correlated with both age and D-dimer.

Levels of significantly altered CTSL1, MCP-3 and IL-6 between symptomatic patients requiring and not requiring O_2_ support were positively correlated with age and LDH levels, while myoglobin positively correlated with both age and D-dimer. REN positively correlated with only D-dimer (Fig. 4B). Interestingly, appetite-stimulator agouti-related protein (AGRP) negatively correlated with D-dimer but not significantly with BMI. PTX3, on the other hand, was positively correlated with not only D-dimer but also with BMI.

### Large EVs from critically ill COVID-19 patients augment pulmonary microvascular endothelial injury

We next tested the effect of plasma-derived LEVs on the apoptosis and survival of pulmonary endothelial cells. An addition of EVs isolated from Severe group patient plasma to HPMEC resulted in significantly increased caspase 3/7 activity when compared to the treatment of cells with EVs from Asymptomatic group. Increased trend in caspase 3/7 activity was also observed in HPMECs treated with EVs from Moderate group COVID-positive patients, however this effect was not significant when compared with Asymptomatic group (Fig. 5A). Further, we checked the cell viability of HPMECs 48h after the addition of EVs and observed a decrease in cell survival in the Severe group when compared to the Asymptomatic group (Fig. 5B).

**Figure 5:**
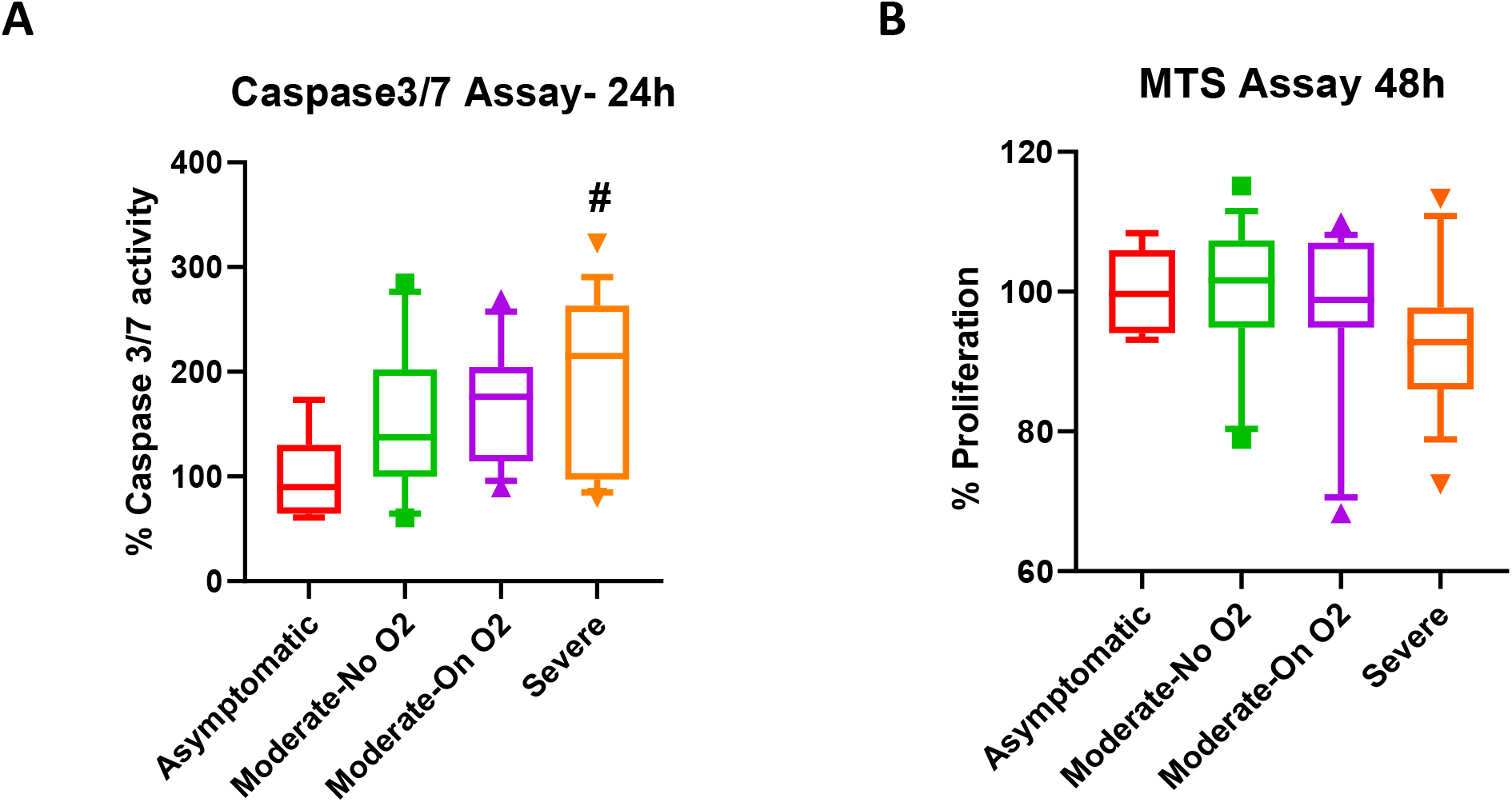
Circulating large EVs from COVID-19 patients promote endothelial dysfunction in the order of disease severity. HPMECs were plated in 96 well plate, followed by addition of µg of LEVs from asymptomatic (n=8), Moderate-No O2 (n=15), Moderate-On O2 (n=15), Severe (n=15). **(A)** Caspase 3/7 assay and **(B)** MTS assay was later performed after 24h and 48h post-treatment, respectively. # p<0.05 vs. Asymptomatic.

## DISCUSSION

In this study, we compared the inflammatory and cardiovascular disease-related protein cargo of EVs from the plasma of COVID-19 patients with different stages of disease severity. We are the first to report significantly enriched LEVs with pro-inflammatory cytokines and chemokines, including proteins from the IL-6 family and TNF superfamily in moderately and critically ill patients with COVID-19. In addition, enhanced levels of proteases, peptidases, and molecules implicated in coagulopathy and endothelial injury were up-regulated in patients with severe COVID-19 disease. Significantly higher EV levels of TF, CD163, and EN-RAGE appear to distinguish severe patients from those in the Moderate-On O_2_ group. Further, we report enhanced apoptosis and attenuated survival of pulmonary arterial endothelial cells on exposure to EVs from patients in the Severe group.

Extracellular vesicles play a key role in the pathogenesis of various diseases, including acute respiratory distress syndrome (ARDS), chronic obstructive pulmonary disease (COPD) pulmonary hypertension (PH), and sepsis(8). Furthermore, EVs released by virus-infected cells can transfer viral proteins, viral receptors, and pro-inflammatory cargo to recipient cells, thereby contributing to the spread of viral infection and worsening tissue injury(9). In a recent study, exosomes released by epithelial cells transduced with lentivirus overexpressing SARS-CoV-2 genes were shown to transfer viral genes to recipient cardiomyocytes leading to an increase in the expression of inflammatory genes(10). Exosomes also have been reported to transfer ACE2 to recipient cells(11). Our OLink analysis indicated presence of ACE2 on circulating LEVs, and, although not significant, ACE2 levels were noted to be higher in Moderate-No O_2_ group patients compared to patients on O_2_ support (data not shown). Negative correlation was observed between ACE2 expression on LEVs and LDH (Cor=-0.8571, P= 0.0238) in Severe group patients without any correlation with age or D-dimer.

Over time, it has become evident that the profound immune response to SARS-CoV-2 is a primary driver of illness severity. Because SARS-CoV-2 enters cells by binding to ACE2 receptors, the renin-angiotensin-aldoserone (RAAS) system and its ties to various inflammatory cascades has been implicated in COVID-19 pathobiology (12). Previously published studies pertaining to the SARS-CoV epidemics have included application of ultra-high-throughput serum and plasma proteomics to identify circulating protein biomarkers of COVID-19 disease(13).We found that EVs from patients infected with COVID-19 carry higher levels of cytokines, including members from the TNF superfamily and IL-6 family, chemokines, such as MCP-1 and CXCL16, and proteases and peptidases, like cathepsin L1, according to the severity of the illness.

Recently, single-cell RNA sequencing using peripheral blood mononuclear cells (PBMCs) from healthy controls and patients with mild or severe COVID-19 or influenza revealed up-regulation of TNF/IL-1P signaling in COVID-19-infected individuals compared to patients with influenza(14). Along these lines, we report elevated levels of multiple members of the TNF superfamily and their receptors in LEVs including TRAIL-R2, TRAIL-R3, TNFRSF10A/DR4, TNFSF13B, TNFSF14,

OPG (TNFRSF11B) and TRAP. Each of these proteins was at a significantly higher level in patients on oxygen support compared to those that were not on oxygen. Whereas, expression of TRANCE, a known suppressor of TNF-α and IL-6 (also known as receptor activator of nuclear factor kappa-B ligand RANKL) was down-regulated in the Moderate and Severe groups compared to asymptomatic patients.

Interleukin-6, known to influence osteoclast and osteoblast activity through RANK/RANKL/OPG interactions has been implicated in the cytokine storm associated with ARDS, septic shock, and COVID-19 disease(15-17).Further, IL-6 is capable of activating nuclear factor kappa-light-chain-enhancer of activated B cells (NF-κB) signaling pathways, which can lead to robust innate and adaptive immune response and promote cellular crosstalk between immune and stromal cells.^18^ Here we found that LEVs from the plasma of patients with COVID-19 contain key factors involved in regulating IL-6 family signaling, including IL-6, IL-6 RA, OSM, IL-27 and LIF-R. OSM induces IL-6, maintains hematopoietic homeostasis, and, in synergy with TNF and IL-1 p, also promotes increased cytokine/chemokine expression and tissue destruction(18).While LEV IL-6 levels significantly correlated with both age and LDH, a circulating marker of tissue injury, positive correlation of IL6-RA and negative correlation of LIF-R on LEVs was observed only with LDH levels.

In addition to IL-6 family cytokines, we found that LEVs from patients with severe COVID-19 contain significantly higher levels of IL-8 compared to patients without severe disease which correlated with LDH and not age. IL-18 levels were also elevated in LEVs from patients with both moderate and severe infection. Moreover, LEV levels of IL-18R1 mirrored those of IL-18 levels, both increasing with disease severity. Accordingly, circulating levels of IL-18 and IL-8 have previously been reported to increase in critically ill patients with COVID-19 (14). While delayed or impaired type I IFN signaling has been implicated in severe COVID-19 infection, the role of the type II IFN signaling pathway is less clear. IFN-γ levels may be mildly elevated in non-severe infection, but levels from the serum and bronchoalveolar lavage fluid of patients with severe COVID-19 have not been found to be significantly elevated in published findings (19).In fact, the increased IL6/IFN-γ ratio has been proposed as a predictor of disease severity (20).Our LEV analysis supports these findings as levels of IFN-γ and IL-12B, an inducer of IFN-γ, were both higher in the Moderate-On O_2_ group than in the Severe group.

Further, LEV levels of the pro-inflammatory RAGE and EN-RAGE were observed to rise with clinical progression of COVID-19. Importantly, EN-RAGE was significantly higher in the Severe group compared to the Moderate-On O_2_ group and correlated with age, a factor, which, itself, is associated with disease severity. These proteins are implicated in lung fibrosis and remodeling and sepsis-related ARDS and have been reported to enhance in COVID-19 infection (15). Recently, anti-inflammatory HGF has been identified as the most significant and specific cytokine biomarker to distinguish severe from non-severe cases of COVID-19 (22).In line with these findings, we also observed increased EV levels of HGF that corresponded with infection severity and correlated positively with LDH levels but not with age.

Monocytes and macrophages appear to play an important role in COVID-19 immunopathology. Monocyte differentiation antigen CD14 and soluble CD163, a marker for monocyte activation, have been shown to be elevated in patients with COVID-19 and potential biomarkers of disease progression (13).Corroborating the pivotal role of monocyte/macrophages in disease severity, we observed significantly higher levels of CD163 in LEVs from severe patients compared to Moderate-On O_2_ group individuals. These increased levels of CD163 positively correlated with D-dimer levels with no correlation observed with age. Furthermore, macrophage CSF-1 was also highly expressed in LEVs from symptomatic COVID-19 patients. It has been reported that EVs derived from macrophages are rich in inflammatory markers such as cytokines, chemokines, and proteases, which could cause destruction of alveolar walls as well as pulmonary vasculature(24-27).Post-mortem analysis of COVID-19 lung sections showed infiltration of mononuclear cells and neutrophils on H&E staining, as well as endothelial cell apoptosis by caspase-3 staining(28).We show here that exposure of HPMECs to LEVs isolated from COVID-19 patients, results in apoptosis and decreased survival of these cells. Based on these findings and positive correlation of CD163 levels with the severity of disease, we speculate a pivotal role of macrophage-derived CD163-positive LEVs in COVID-19 related lung injury.

Since the start of the pandemic, D-dimer levels have been used as a marker of disease progression and poor outcomes (30). Microvascular thrombosis similar to that seen in sepsis-induced coagulopathy and disseminated intravascular coagulopathy has been described and implicated in COVID-19 (31). Together with the vasoconstriction and stagnant blood flow induced by elevated levels of Angiotensin II, inflammation and endothelial injury set the stage for coagulopathy. EVs have been shown to release or present TF and pro-coagulant phospholipids on their surface promoting clot formation and to accelerate fibrin polymerization (32). We also found significantly elevated levels of TF, t-PA, and vWF in LEVs from patients with severe COVID-19 compared to healthy controls. Elevated levels of tPA, a protease responsible for the conversion of plasminogen to plasmin, were previously reported in patients infected with SARS-CoV-1(34). More recently, vWF and Factor VIII, both released from injured endothelial cells, were also shown to be increased in COVID-19 (35). In addition, we observed reduced levels of ADAMTS13 in EVs from severe COVID-19 patients compared to healthy controls and it was negatively correlated with D-dimer levels and age. ADAMTS13 is known to regulate micro-vascular thrombi formation by cleaving ultra-high-molecular-weight vWF multimers and a similar secondary deficiency in ADAMTS13 has been described in other systemic illnesses (34).

It is important to note that in our study population and in COVID-19-infected individuals globally, hypertension is one of the most common preexisting comorbid conditions (37). Plasma renin, a hormone responsible for initiating the RAAS cascade and regulating blood pressure, was elevated in EVs from COVID-19 patients with severe disease compared to healthy controls. Renin also positively correlated with D-dimer levels when comparing EVs from hypoxic patients to those not requiring oxygen. PRSS8, another protein involved in cardiovascular disease, was the protein most up-regulated in LEVs from severe COVID-19 infection compared to healthy individuals. We also observed elevated LEV levels of several peptidases and proteases implicated in vascular remodeling, including CTSL1, MMP9, CPA1 and CPB1. Myocardial injury, whether through direct viral infection or induced by cytokine storm or hypoxia-induced apoptosis, has been seen in 7% of all infected patients and 22% of those admitted to an ICU (37).We noted elevated levels of myoglobin, a biomarker of cardiac muscle injury, in LEVs from patients with moderate or severe COVID-19.

While our results show multiple, promising insights into the pathophysiology of COVID-19 infection and the potential role of circulating EVS in the related immune response, our study does have some limitations. We were limited in the number of healthy controls available to compare against our four infected groups. The healthy samples we were able to obtain were not drawn under the same conditions as those enrolled in the biorepository, such as a fasting condition or time of day. We also lacked an uninfected control group with similar comorbidities, including hypertension and diabetes. Overall, our samples sizes were relatively small, and future analysis using larger cohorts is warranted. Furthermore, in addition to LEVs, significantly increased circulating small EVs in severe COVID-19 patients are also believed to play an important role in pathogenesis and analysis of SEV cargo is currently part of our ongoing studies.

In conclusion, this unique study contributes to the expanding knowledge of the pathophysiology of this highly infectious and devastating disease. We suggest that large extracellular vesicles play a significant role in COVID-19 patients by contributing to the augmented pro-inflammatory response, endothelial dysfunction and micro-thrombosis observed in patients who have more severe disease. Perhaps this knowledge around EVs will contribute to determining potential biomarkers for poorer outcomes in COVID-19 infection and offer a novel way to approach therapies to treat this disease and other viral-related illnesses.

## Data Availability

The datasets generated during and/or analyzed during the current study are available from the corresponding author on reasonable request.

## ACKNOWLEDGEMENTS

We acknowledge Ashok Kumar, Huiqing Yin-DeClue and Ling Chen, Department of Internal Medicine, KUMC for their help in processing blood specimens. We thank Luigi Boccardi, Department of Internal Medicine, KUMC for his assistance in the preparation of IRB protocols, consent forms and other regulatory documents for COVID-19 bio-repository. We also acknowledge Electron Microscope Research Laboratory (NIH/NIGMS COBRE grant P20GM104936 and NIH grant 1S10RR027564) for TEM analysis.

